# Evaluating and Reducing Subgroup Disparity in AI Models: An Analysis of Pediatric COVID-19 Test Outcomes

**DOI:** 10.1101/2024.09.18.24313889

**Authors:** Alexander Libin, Jonah T. Treitler, Tadas Vasaitis, Yijun Shao

## Abstract

Artificial Intelligence (AI) fairness in healthcare settings has attracted significant attention due to the concerns to propagate existing health disparities. Despite ongoing research, the frequency and extent of subgroup fairness have not been sufficiently studied. In this study, we extracted a nationally representative pediatric dataset (ages 0-17, n=9,935) from the US National Health Interview Survey (NHIS) concerning COVID-19 test outcomes. For subgroup disparity assessment, we trained 50 models using five machine learning algorithms. We assessed the models’ area under the curve (AUC) on 12 small (<15% of the total n) subgroups defined using social economic factors versus the on the overall population. Our results show that subgroup disparities were prevalent (50.7%) in the models. Subgroup AUCs were generally lower, with a mean difference of 0.01, ranging from -0.29 to +0.41. Notably, the disparities were not always statistically significant, with four out of 12 subgroups having statistically significant disparities across models. Additionally, we explored the efficacy of synthetic data in mitigating identified disparities. The introduction of synthetic data enhanced subgroup disparity in 57.7% of the models. The mean AUC disparities for models with synthetic data decreased on average by 0.03 via resampling and 0.04 via generative adverbial network methods.

## 1. Introduction

Artificial Intelligence (AI) models are increasingly used in healthcare. Many shareholders have high expectations that AI would revolutionize healthcare and healthcare research, and there have been some early signs of success. (1, 2) One example is using machine learning (ML) methods to detect diabetic retinopathy. Several classification models trained using different datasets have achieved good (>90%) sensitivity, specificity, or AUC. (3) The recent development of significant language model-based generative AI has further increased the enthusiasm. Since the introduction of Generative Pre-Trained Transformers (GPT), several studies have shown that it was able to generate answers that have accuracy to pass the United States Medical Licensing Exam. (4-6)

Meanwhile, serious concerns have been raised that AI may perpetuate health disparity (7-9). Real-world data are subjected to biases as they may contain missing entries, suffer from class imbalance, be affected by sampling biases, and not have been collected for modeling. AI models that are trained on biased information thus can yield biased results. Sampling bias, for example, can be introduced in a dataset when only some with a particular characteristic are recruited into a study while others are systematically excluded. (10) In this case, an AI algorithm trained on the data can inadvertently make a poor prediction for the underrepresented subgroup and further exasperate the inequalities within the health care system. Commonly used fairness metrics include the disparity between subgroups regarding AUC (area under curve, measures overall distinguishing power), accuracy, and true positive and false positive rates. (11) Some studies assessed the fairness of health risk prediction models focusing on specific vulnerable populations such as African Americans. (12) Other studies developed mitigation strategies, including generating and adding synthetic data to an existing dataset. (13, 14)

Many knowledge gaps exist despite the strong interest in AI fairness in the biomedical domain. Literature has shown that some trained models perform better in one subgroup than another, but it is unclear how often and to what extent subgroup disparity exists in health risk prediction. Similarly, more experiments are needed to determine how often and to what extent the new mitigation strategies, such as synthetic data generation, can enhance disparity.

In this study, we use pediatric COVID-19 risk prediction as the use case. Since 2020, COVID-19 has affected millions of children. Children, including teenagers, tend to have milder COVID-19 symptoms. Nevertheless, COVID-19 became one of the leading causes of death among those between 0-19 years of age. (15) Children, like adults, suffer from health disparities. For example, most children are healthy, but a small subpopulation has underlying conditions (e.g. diabetes) and is thus particularly at risk for (severe) COVID-19. {Bishop, 2024 #1290}

While researchers have discussed the fairness and biases of AI pediatric health risk prediction models, (25) among the studies that trained ML models for pediatric COVID-19, (26, 27) few assessed the subgroup disparity of the ML models (i.e., the disparity of model performance between different subgroups of the population). Our study is novel in that we sought to assess the prevalence of subgroup disparity when modeling COVID-19 test outcomes in children and test if the disparities are statistically significant. In addition, we selected two subgroups with statistically significant disparities to test the effectiveness of using synthetic training data to reduce subgroup disparities.

## 2. Methods

### 2.1 Dataset

We obtained a set of the US National Health Interview Survey (NHIS) data (2020-2022) from the IPUMS (16) and identified a cohort of children (ages 0-17). (Table 1) NHIS is a nationally representative sample. It is “the principal source of information on the health of the civilian noninstitutionalized population of the United States and is one of the major data collection programs of the National Center for Health Statistics (NCHS), which is part of the Centers for Disease Control and Prevention (CDC).”(17) Comparing to electronic medical record data, NHIS has more detailed information on social determinants health such as citizenship and health insurance status which we used to define subgroups.

**Table 1.** Demographics and other characteristics of the NHIS study cohort. NHIS has detailed demographics with limited clinical data.

|  | **N=9935** |  |
| --- | --- | --- |
|  | **Mean/N** | **Std/%** |
| **Age** | 9.1 | 5.3 |
| **Gender** |  |  |
| *Female* | 4811 | 48.2% |
| *Male* | 5124 | 51.6% |
| **Race** |  |  |
| *White* | 7151 | 72.0% |
| *Black* | 1159 | 11.7% |
| *Asian* | 722 | 7.30% |
| *Mixed Race* | 649 | 6.50% |
| *Alaska Native/Native American* | 250 | 2.50% |
| **Ethnicity** |  |  |
| *Non-Hispanic/Spanish Origin* | 8029 | 80.8% |
| *Mexican* | 1147 | 11.5% |
| *Other Hispanic* | 759 | 7.60% |
| **Other characteristics** |  |  |
| *US Citizen* | 9701 | 97.6% |
| *Receives Food Stamps* | 1879 | 18.9% |
| *Free or Reduced Lunch* | 4338 | 43.7% |
| *Have Insurance Coverage* | 9500 | 95.6% |
| *Excellent Health* | 6451 | 64.9% |
| *Very Good Health* | 2261 | 22.8% |
| *Good Health* | 983 | 9.9% |
| *Fair Health* | 205 | 2.1% |
| *Poor Health* | 35 | 0.4% |
| *Received Flu Vaccine* | 4903 | 49.4% |
| *ADD* | 1026 | 10.3% |
| *Asthma* | 1094 | 11.0% |
| *Positive COVID-19 test* | 2612 | 26.3% |

NHIS data contain thousands of variables, though most are not included in every survey for every age group. In the surveys between 2020 and 2022, NHIS added a set of questions related to COVID-19. For the ML experiments described here, COVID-19 test result (positive vs negative) was chosen as the outcome variable. Some other COVID-19 outcome questions were only included in the surveys for one or two quarters, limited to adults, or had very few positive answers. In pre-processing, we treated various “unknown” answers (e.g., “unknown-refused” or “unknown-don’t know”) as missing values. Variables with high amounts of missing values were removed from consideration as predictive features. We also focused on selecting variables discussed as risk factors in the COVID-19 literature. (18-24) The demographics and other characteristics of the study sample are shown in Table 1.

### 2.2 Subgroup disparity assessment

For the subgroup disparity assessment, we selected five ML algorithms (gradient boosting machine (GBM), neural network (NN), penalized logistic regression (PLR), support vector machine with linear kernel (SVM), and k nearest neighbor (KNN)), that are very different in their modeling approach. For example, PLR and SVM with linear kernels yield linear models, while GMB, NN, and KNN can model complex, non-linear relationships: PLR is a variant of logistic regression that incorporates regularization techniques to prevent overfitting and improve generalization by adding penalty terms to the cost function. SVM is an algorithm that finds a hyperplane that best separates different classes in the feature space. GBM is an ensemble algorithm that combines multiple decision trees. NN models consist of interconnected nodes intended to mimic human neurons organized in layers. KNN is a simple algorithm that predicts the class of a new data point based on the majority class or average value of its k-nearest neighbors in the feature space.

Before fitting each model, the data was randomly split into 90% training and 10%. We then fitted the models on the training data with parameter selection using a five-fold cross-validation. The trained model was then applied to the testing data. The training and testing were repeated 10 times per ML algorithm. This yielded 50 different models for the evaluation of fairness.

To assess the subgroup disparity, 12 small (<15% of the total n) subgroups, including 6 racial and ethnic minorities, were selected. For each model, we calculated and compared the AUCs (area under the curve, measuring overall distinguishing power) of the overall population and each subgroup in the testing data. Theoretically, any difference in model performance between two subgroups can be called subgroup disparity. In this study, we defined subgroup disparity as a model with *worse* AUC on a small subgroup than the overall population.

We calculated the prevalence and magnitude of subgroup disparity measured as AUC differences. Because the AUCs are affected by the ML algorithm and the partition of training and testing data, we used paired t-tests to assess the statistical significance of the subgroup disparity between a subgroup and the overall population.

### 2.3 Use synthetic data to reduce subgroup disparity

To reduce subgroup disparity, we tested the use of synthetic data. Two subgroups with AUCs statistically significantly lower than the general population were selected. Two methods, oversampling and generative adversarial network (GAN), were used to generate synthetic data. For oversampling, we used the built-in resample function in R. A GAN learns to generate synthetic data by learning the joint distribution in the actual data through two neural networks, one function as the generator and the other as the discriminator. We used the RGAN package in R for this experiment. (28)

We set the synthetic data sample size to the sample size of the two selected subgroups in the training data. In other words, we doubled the sample size of the subgroups by adding synthetic data. We fitted NN models with and without synthetic data but evaluated them using the same testing data.

The training and testing were repeated 10 times with different random 90:10 split of training and testing data. The AUC differences in models fitted with and without synthetic data were compared. (Fig. 1) Given the goal of reducing subgroup disparity, the evaluation focuses on models with AUC disparities associated with the two selected subgroups. This experiment used NN as the ML algorithm because it had one of the best mean AUCs.

**Figure 1.**
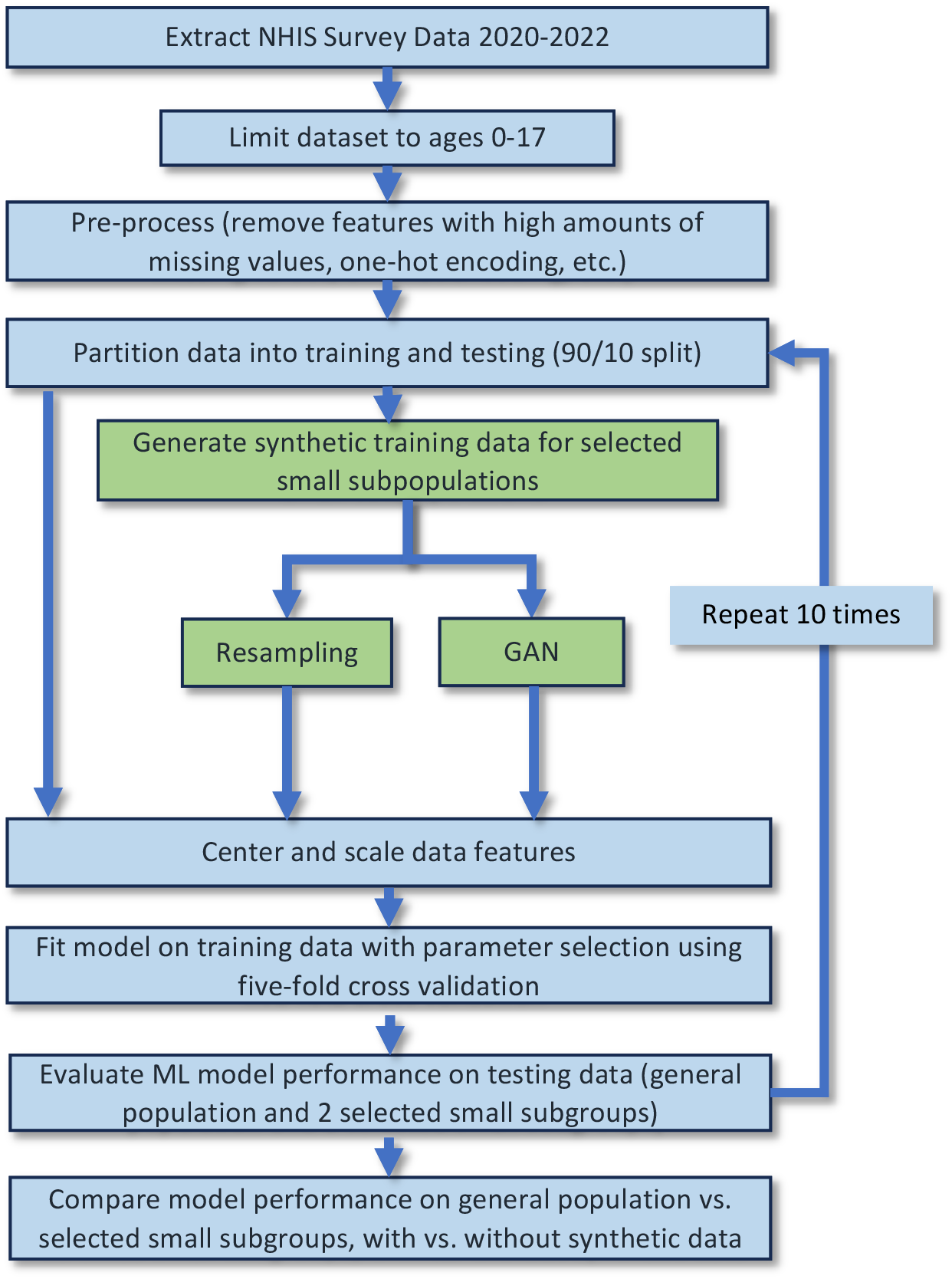
Experimental design for testing the effect of increasing the sample size of selected small subgroups using synthetic data.

## 3. Results

In the subgroup disparity assessment, we compared the AUCs of 50 models (5 algorithms with 10 different data partitions) in the 12 small subgroups vs those in the general population. (Table 2) Our results showed that 50.7% of the time, the AUC of a small subgroup was lower than that of the general, suggesting that subgroup disparity is prevalent. There is, however, a large variation in the AUC differences: Mean: 0.01, Min: -0.29, Max: 0.41, SD: 0.09. (A positive difference indicates an AUC disparity.)

**Table 2.** Average AUC and AUC disparity by ML method. A positive number indicates disparity, i.e., the ML model(s) performed worse on the subgroup than the overall population. KNN: K nearest neighbor; GBM: Gradient boost machine; PLR: Penalized logistic regression; NNET: neural network.

| ML Method | Mean AUC on overall population (SD) | AUC Disparity (SD) |
| --- | --- | --- |
| *KNN* | 0.58(0.01) | -0.01(0.10) |
| *GBM* | 0.63(0.01) | 0.00(0.09) |
| *SVMLinear* | 0.63(0.03) | 0.02(0.10) |
| *PLR* | 0.64(0.01) | 0.03(0.09) |
| *NN* | 0.64(0.01) | 0.00(0.09) |

The paired-t tests found that in four of the 12 subgroups (Non-Citizen, Good Health, Black, and Native American), the AUC disparities were statistically significant. (Table 3) A few of the other subgroups, on the other hand, has better AUCs than the general population. We need to note that on small sample sizes, the assessment of AUC is not as reliable as on larger sample sizes.

**Table 3.** The mean and AUC of each subgroup. Four subgroups had statistically significantly worse AUC than the general population. *Since most survey participants have excellent and very good health, good health is a subgroup with less optimal outcomes.

| Subgroup | Mean AUC (SD) | Mean AUC difference | P value |
| --- | --- | --- | --- |
| Non-Citizen | 0.58 (0.13) | 0.05 | 0.02 |
| Good Health* | 0.60 (0.06) | 0.02 | <0.01 |
| Black | 0.60 (0.06) | 0.02 | <0.01 |
| Native American | 0.67 (0.14) | 0.05 | <0.01 |
| Very Good Health | 0.63(0.13) | -0.01 | 0.72 |
| No Insurance Coverage | 0.62(0.12) | 0.01 | 0.74 |
| Mexican | 0.61(0.07) | 0.02 | 0.08 |
| Other Hispanic | 0.61(0.07) | 0.01 | 0.11 |
| ADD | 0.63(0.07) | -0.01 | 0.25 |
| Asthma | 0.64(0.05) | -0.02 | <0.01 |
| Asian | 0.65(0.07) | -0.02 | 0.01 |
| Other Race and Multiple Race | 0.57(0.09) | -0.02 | 0.22 |

In the synthetic data experiment, we focused only on models that showed disparity in regard to the two selected subgroups. (Figure 2) The addition of synthetic data reduced the subgroup disparity in 57.7% of these models: oversampling reduced AUC disparity by 63.6%, and GAN reduced the AUC disparity by 53.3%. In the cases where AUC disparity was reduced, the disparity was decreased by an average of 0.03 from 0.08 to 0.05, with the average reduction associated with resampling being 0.03 and the average reduction associated with GAN being 0.04. As shown in Figure 3, the disparity results from a combination of the increase of the subgroup AUC and the decrease in the overall AUC.

**Figure 2.**
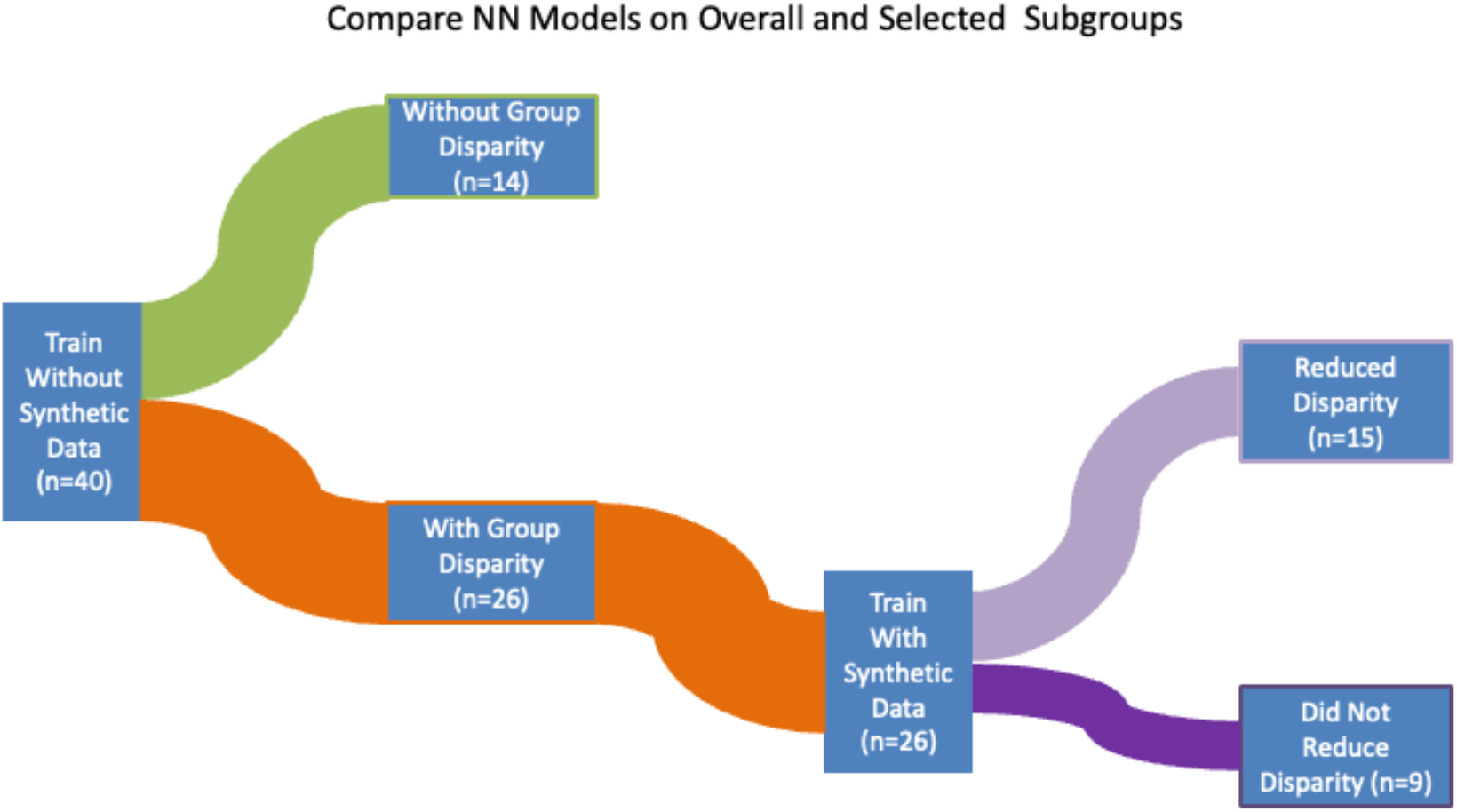
The synthetic data experiment showed that most original models had subgroup disparities, and the addition of synthetic data to model training led to reduced disparities in most cases where subgroup disparities were observed.

**Figure 3.**
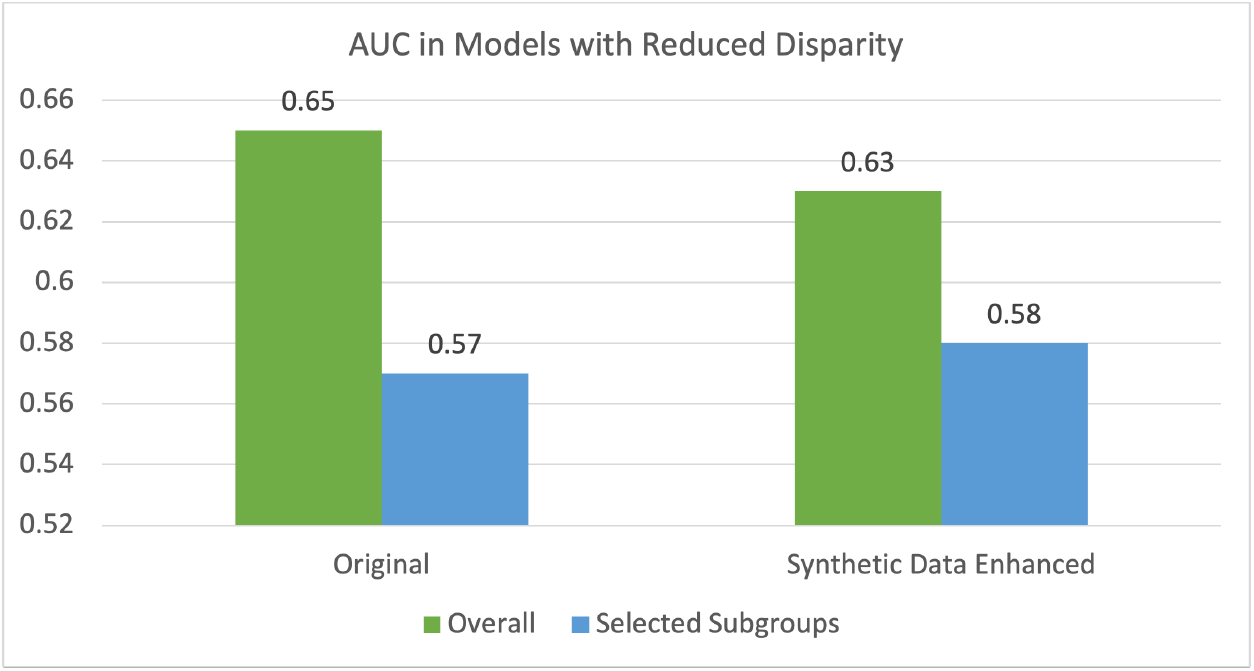
The mean AUCs of the selected subgroups vs. overall subgroup in models with subgroup disparity were reduced by adding synthetic data.

## 4. Discussion

### 4.1. Findings

Most of the prior publications that measured subgroup disparity of AI prediction in the biomedical domain focused on one or two models and reported findings of disparities. In our experiments, using five different machine learning algorithms, random data partitions, and twelve subgroups, we found that subgroup disparity is prevalent (50.7%) when modeling COVID-19 outcomes in children (0-17 years old). In other words, MLmodels often performs worse on a small subgroup than on the overall population.

However, the high prevalence of subgroup disparity is partly due to random effects associated with partitioning training and testing data and the use of random seeds in algorithms. We observed that the mean difference between the subgroup AUCs and overall AUCs is only 0.01, with a minimum of -0.29, a maximum of 0.41, and a standard deviation of 0.09. This suggests that the average AUC of the small subgroups was only slightly lower than that of the overall population. Sometimes, the small subgroups’ AUC are higher and sometimes lower than the overall population AUC.

The subgroup disparities are not always statistically significant. We found that four out of the twelve subgroups had statistically significantly lower AUCs than the general population in the paired t-test. This suggests that these four subgroups had different predictor-outcome relationships from the general population.

Synthetic data has been reported to boost AI model performance and used as a bias-mitigation strategy. We experimented with generating synthetic data through oversampling and GAN to reduce the prediction disparities in two subgroups that with statistically significant disparities. Our results suggest that the oversampling and GAN methods can but do not always lead to lower subgroup disparity.

### 4.2. Implications

When training for healthcare risk prediction models, researchers and developers often optimize for the overall performance, such as AUC, accuracy, and positive and negative predictive values. As our analyses suggest, the subgroup disparity is prevalent. The mean disparity in our study was small (0.01) but the maximum disparity was very large (0.41). Selecting a model with low average or low maximum subgroup disparity would be desirable when choosing a risk prediction model. A model, however, cannot be expected to achieve the exact performance in every subgroup since subgroups often have somewhat different characteristics and may have somewhat different underlying relationships between predictors and outcomes.

Statistically significant subgroup disparities may be caused by several factor. (29) In our study, for example, the non-citizens and Native Americans have a very small smple size. It has been reported that native youths suffered the most considerable loss rate of caregivers. (30) Hill et al discussed immigration status as a health care barrier in the US during COVID-19 in their 2021 paper. (31) To improve the model performance on these subgroups, we do not only need to increase the sample size but should also examine confounders (e.g., body mass index, diabetes diagnosis, length in the US for immigrants) that were not captured by the NHIS data on children.

While it is possible for a given model to perform better on specific smaller subgroups (as shown in our experiments), ML algorithms are typically designed to optimize their performance for the majority subgroup. As a result, de-biasing strategies have been designed to boost modeling training on the small subgroups through weighing, oversampling, and synthetic data generation. (32) Our results show that over-sampling and GAN-based synthetic data generation could sometimes reduce the bias. Even when the reduction was achieved, they do not always completely eliminate the disparities.

### 4.3. Limitations

One limitation is that we only measure AUC, while other ML performance measures include sensitivity, specificity, and accuracy may be more relevant in some clinical tasks. We did not choose those other metrics in our experiment because they are threshold-specific. However, based on the use case, a threshold-specific metric may be more appropriate.

Another limitation is the sample size. While a sample of almost 10,000 patients is not too small for ML in general, GAN typically requires a larger amount data to train. The size of the small subgroups may be simply too small to take advantage of GAN’s capabilities. This is also why we did not select non-citizen and native American subgroups for synthetic data generation.

A very challenging issue for most existing de-basing algorithms is the multi-minority status. Re-weighting methods, for example, are usually designed to enhance the performance of one sensitive variable or subgroup. While we examined multiple sensitive variables in this study, we did not consider multi-minority status.

In addition, there is sometimes a tradeoff between overall group performance and subgroup performance. When we added the synthetic data for model training, the average overall AUC was reduced by 0.02, while the disparity in AUC was reduced by 0.04. In other words, the reduction in disparity came with a cost in overall performance. One way to address this issue may be to create a subgroup-specific model that optimize the subgroup performance through transfer learning. (33)

### 4.4. Future work

In future studies, we would like to test other bias mitigation methods on larger samples. We also plan to study the subpopulation with multi-minority status, which may be the most vulnerable patient subgroup.

## 5. Conclusions

We assessed AI fairness in the context of pediatric COVID-19 test result outcome prediction as well as experimented with the use of synthetic data to reduce subgroup disparities. Our results suggest that subgroup disparity is prevalent in ML models, though often not statistically significant, and synthetic data can sometimes enhance subgroup parity.

## Data Availability

The datasets generated during and/or analyzed during the current study are publicly available.

https://www.cdc.gov/nchs/nhis/data-questionnaires-documentation.htm

## Author Contributions

Conceptualization, Alexander Libin and Yijun Shao; Data curation, Jonah Treitler; Formal analysis, Jonah Treitler and Yijun Shao; Funding acquisition, Tadas Vasaitis; Methodology, Yijun Shao; Project administration, Alexander Libin; Resources, Alexander Libin and Yijun Shao; Supervision, Alexander Libin; Visualization, Jonah Treitler and Yijun Shao; Writing – original draft, Alexander Libin and Jonah Treitler; Writing – review & editing, Alexander Libin and Tadas Vasaitis.

## Institutional Review Board Statement

The study was conducted in accordance with the Declaration of Helsinki. No IRB approval is needed because the dataset being used is publicly available.

## Data Availability Statement

The datasets generated during and/or analyzed during the current study are publicly available.

## Acknowledgments

We thank the NHIS and the IPUMS for making their data available to us. This research was, in part, funded by the National Institutes of Health (NIH) Agreement No. 1OT2OD032581. The views and conclusions contained in this document are those of the authors and should not be interpreted as representing the official policies, either expressed or implied, of the NIH. Conceptual approach reported in this publication was supported in part by the National Center For Advancing Translational Sciences of the National Institutes of Health under Award Number UL1TR001409. The content is solely the responsibility of the authors and does not necessarily represent the official views of the National Institutes of Health.

